# Statistical features of complex systems in use of pre-hospital emergency services: a linked database study

**DOI:** 10.64898/2026.05.18.26352011

**Authors:** James Cussens, Kelly - Ly Phuong Thuy, Emily V Chambers, Annabel Crum, Chris Burton

**Affiliations:** Sheffield Centre for Health & Related Research, University of Sheffield, Regent Court, 30 Regent St, Sheffield S1 4DA

**Keywords:** Emergency care, prehospital care, high intensity use, frequent attendance, complex systems, power law

## Abstract

**Background:** High Intensity Use of urgent medical services by patients is widely recognised in urgent and emergency care. Studies of high intensity use of the emergency department have consistently shown features of complex systems behaviour in addition to highly heterogeneous individual patient characteristics. There have been no comparable studies of prehospital care use.

**Methods:** We examined the use of prehospital urgent and emergency services (NHS 111 and ambulance dispatch) using routinely collected data from regional service in the UK (population 5 million). We used a complex systems perspective, to examine (1) distribution of contacts per individual; (2) the temporal stability of service use by individuals and at the whole-system level (3) the distribution of bursts of contacts.

**Results:** We analysed data from 847555 individuals who contacted NHS111 and 389550 who contacted the ambulance dispatch service. 35120 (4.2%) individuals who contacted NHS111 had 5 or more contacts with the service over the two-year period and accounted for 290625 (20.1%) of contacts. 16755 (4.3%) individuals had 5 or more ambulance dispatch contact days and accounted for 169085 (25.8%) of contacts. The distribution of contacts per individual showed a monotonic distribution between 5 and over 100 contacts that was heavy tailed and compatible with a power law distribution. At any level of use, patients with one or more mental health related contacts had a greater likelihood of further contact than those without.

**Conclusion:** Prehospital emergency service use shows multiple statistical features typical of a complex system. Interventions to manage demand need to consider both individual high intensity users (particularly in relation to their mental health) and the behaviour of the whole system.

## Introduction

High intensity users (people who access healthcare services more frequently than the majority of the population) are seen in all urgent and emergency care settings(1-3). The common reasons for high intensity users (HIUs) accessing services include severe or unstable disease needing treatment(3), intrusive and distressing symptoms that are disproportionate to underlying physical disease(4, 5), recurrent mental health difficulties, (6) and complex social circumstances (7) particularly when no other immediate support is available (8). While these reasons can exist independently of each other, they commonly overlap.

HIUs are consistently found in emergency departments (ED) and GP out of hours service (GPOOH)(1). They are also increasingly recognised in pre-hospital urgent and emergency services including NHS 111 and 999(3, 9). In UK prehospital urgent and emergency services, high intensity use (also known as frequent calling) is defined as 5 or more contacts in a month or 12 contacts over a three month period(3, 10). This threshold is considerably higher than equivalent thresholds for high intensity use of the ED (typically 5 attendances in a 12 month period), suggesting that many calls from HIUs do not result in ED attendance. Since 2014, Ambulance Services in England have been required to provide specific services for HIUs. While these vary, they typically involve case management(11, 12). This appears particularly suited to HIUs with complex social problems, especially when these feed into recurrent mental health difficulties. The effectiveness of case management for HIUs of ambulance services has recently been evaluated: although there were instances of case management being apparently successful for some individuals, there was no impact on patterns of high use.(13) In part, this may be a consequence of the heterogeneity of the high user population: it highlights the need for more specific interventions targeted at particular patterns of high use.

While conventional approaches to understanding the problem of high use have focused on characteristics of individual patients, there is increasing interest in taking a complex systems approach to understanding the challenges of urgent healthcare delivery (14-16). While typically such work is qualitative, we have previously examined high intensity use of ED and primary care data using a quantitative approach. (1, 17, 18). These studies have consistently found statistical features typical of complex systems: specifically (i) observed distributions of attendances per person show good fit with well-defined heavy tailed distributions including the inverse power law (1, 18) which is indicative of a complex system (19) (ii) shown high instability of individual behaviour over time but high stability of system behaviour across subgroups (1, 18); (iii) identified patterns of bursts of attendance as a potential generative process (1, 20). While this formal complex systems approach to understanding attendance is not widely used, it is compatible with qualitative knowledge of how patients use services (11, 21) and with a broader social model of how services are used. (22)

We aimed to analyse patterns of high intensity use of ambulance services, with a particular focus on adults aged 18-65 in order to (i) identify patterns of high intensity use of ambulance services and the factors associated with them (ii) Describe how these patterns vary within individuals over time (iii) Identify patterns of symptoms which may be indicative of sustained high use, or suitable for interventions.

## Methods

### Study design

This was a retrospective cohort study of routinely collected ambulance service data linked at the (anonymised) individual patient level.

### Data source

We used a bespoke dataset created from the CUREd+ database, a real-world linked research platform for analyzing the urgent and emergency care system (23). For this study we examined routinely collected data from the Yorkshire Ambulance Service (YAS) which provides pre-hospital urgent and emergency care for the Yorkshire region of England comprising a population of approximately 5 million with a mix of urban and rural geographies and wide variation in socioeconomic status. YAS provides two services: the NHS111 telephone triage service and the ambulance despatch service. Both services are accessed directly by the public (using the numbers 111 and 999 respectively) and are integrated such that calls to one service can be routed to the other as appropriate. All data was accessed using the secure data environment provided by the University of Sheffield Data Connect service. Ethical approval was granted by the University of Sheffield (Reference Number: 067407).

#### Data specification

Data for the study comprised data relating to contacts with the 111 and 999 services between April 2021 and March 2023. The bespoke dataset was comprised of relevant fields from four datasets, all linked by a common pseudonymised ID: Demographics (age, sex and Index of Multiple Deprivation of first stated home address) ; 111 Calls Dataset (2017-2023); Yorkshire Ambulance Service Computer Aided Dispatch (2017-2023); and Yorkshire Ambulance Service Electronic Patient Record (EPR) (2017-2023).

For this we used the following data: Pseudonymised ID at each contact as a linkage item; age, gender and Index of Multiple Deprivation of home address at the first contact; date & time of each contact; clinical reason for contact () and outcome of each contact (advice, referral to other services, transfer to hospital). We limited data to adults aged 18-65 as including older groups will introduce issues of age-related frailty which is not the focus of this research (our previous work on Emergency Department high intensity use has shown a change in distribution of attendance above the age of 70). The data was supplied for the NHS 111 and ambulance dispatch datasets and comprised details of each contact linking calls for each individual through a pseudonymised ID, which in turn was linked to the individual’s age, sex and Index of Multiple Deprivation (IMD) quintile of their home address (where available). Basic contact data (service, and date) were supplied for all contacts (with only the first call of each day for an individual included). Additional data was supplied for individuals with 5 or more contacts over the two years with the ambulance dispatch service comprising all contacts (when there were multiple contacts in one day) and including the reason for contact including both text options and codes. This threshold was chosen in advance to represent approximately the 5% of highest users of the service.

#### Data preparation

Prior to analysis, data was cleaned and checked to exclude duplicate calls. In particular, the first dataset included only the first contact on any given day. While the second dataset included multiple contacts on the same day and we initially intended to use these, we found a few instances of individuals who made large numbers of calls on the same day. To avoid this influencing results, particularly in relation to reason for contact, we elected to use only the first call of each day for all individuals.

For analysis, we calculated the number of calls per person (111 or ambulance) in the total 24 month period and for each of 3 consecutive 8 month periods. We categorised reason for attendance on ambulance calls based on the dispatch data into 12 categories. This data contained a mix of specific clinical presentations (e.g. suspected stroke), broad categories (e.g. limb trauma), and non-specific codes (e.g. call requested by healthcare professional or call transfer between 111 and ambulance).

## Statistical analysis

### Distributions of contacts per person over the whole period

We hypothesised that the distribution of contacts per person would approximate to a power law. To examine this across the whole prehospital care system we used the dataset comprising all users. To test this we took two approaches: visualisation and statistical fitting to hypothesised distributions. Visualisation was carried out by plotting the conditional cumulative distribution function (which represents the probability that for any number of contacts x, an individual has x or more contacts). When displayed on logarithmic axis an inverse power law displays as a straight line. Statistical fitting was carried out using the poweRlaw package (24) in R4.6 (R Foundation) using a maximum likelihood function and testing fit using the Vuong test comparing the data to 3 distributions: power law (hypothesised), poisson (for discrete events which are randomly distributed), and log-normal (an alternative heavy-tailed distribution).

### Stability of number of contacts by high users

We examined the stability of contacts over time by calculating the number of contacts per individual in three consecutive 8-month periods. We then categorised the number of contacts per period into three or four categories (e.g. 0,1-4,5-8, 9+) and included data only from individuals whose attendance fell in the highest category for at least one of the 8-month periods. We then plotted transitions between categories in each period using alluvial plots.

### Distribution of bursts of contacts

For each individual we sorted their calls in temporal order and calculated the interval between each pair of consecutive calls. We then allocated each calls to a burst, where a burst was defined as a sequence of contacts for which the inter-contact interval is less than or equal to a time interval *t*. We set values of *t* of 7, 10 and 14 days and limited the analysis to individuals with between 5 and 75 contacts to avoid data being either too sparse or too congested. We then aggregated the bursts across all included individuals and plotted the conditional cumulative distribution function. We hypothesised this should follow a power law, indicating that “bursty activity” could be acting as a generative mechanism for patterns of high use (20).

### Factors associated with high use

We examined the relationship of patient characteristics (age, sex and home deprivation index) on high use as both a continuous and categorical outcome. The continuous outcome approach used logarithmic plots split by categories of characteristics (e.g. median split of age). The categorical outcome approach involved binning the number of contacts into a set of categories and using descriptive statistics and logistic regression. We used categories of 1-4, 5-8,9-12, 13-24 and 25+ contacts. In addition to reporting numbers and proportions, we used logistic regression to estimate the effect of these factors on the probability of having at least a target number of contacts given a threshold number of contacts (e.g. the probability of 5 or more contacts given the presence of at least 1 contact or the probability of 8 or more contacts given the presence of at least 5).

We tested whether the reason for contact in calls was related to high use in the ambulance dispatch dataset by creating additional variables relating to whether one or more contacts belonged to a specific category (e.g. neurological, cardiorespiratory, mental). To avoid the influence of individuals having more reasons for contact just by nature of having more contacts, we restricted the presence of categories to those in the contacts before the threshold value (for example: adding a variable “mental reason for contact in first 8 contacts” to the logistic regression model for those with 12 or more contacts having had at least 8).

### Data disclosure control

All analyses were conducted within the DataConnect Secure Data Environment and used all available data for statistical analyses. However in keeping with the disclosure control policy, values in tables and figures were rounded to the nearest 5 and values equal to or less than 7 were suppressed. For the distribution plots this resulted in grouping of points and omitting the extreme tail of the data.

## Results

### Description of cohort

The wide (whole population) dataset comprised 847555 individuals who had a total of 1442640 contact days with NHS111 and 389550 individuals who had a total of 655345 ambulance despatch service contact days. 35120 (4.2%) individuals who contacted NHS111 had 5 or more contacts with the service over the two-year period and accounted for 290625 (20.1%) of contacts. 16755 (4.3%) individuals had 5 or more ambulance dispatch contact days and accounted for 169085 (25.8%) of contacts. Reasons for contact in the ambulance dispatch dataset of high intensity users are summarised in Supplementary Table 1. Medical reasons for contact including cardio-respiratory (18.0%) and neurological (12.8%) were much more common than trauma (5.1%). Mental health related contacts (including self-harm and substance use) accounted for 15.4% of contacts from the high intensity users.

**Table 1.** Characteristics of service users by categories of use.

| Contact days per person | 1-4 | 5-8 | 9-12 | 13-24 | 25+ | Overall |
| --- | --- | --- | --- | --- | --- | --- |
|  | (N=811740) | (N=27285) | (N=5055) | (N=2675) | (N=800) | (N=847555) |
| **IMD Quintile** |  |  |  |  |  |  |
| 1 | 271625 (33.5%) | 12620 (46.3%) | 2465 (48.8%) | 1290 (48.2%) | 350 (43.8%) | 288355 (34.0%) |
| 2 | 147620 (18.2%) | 4775 (17.5%) | 865 (17.1%) | 410 (15.3%) | 130 (16.1%) | 153795 (18.1%) |
| 3 | 129295 (15.9%) | 3260 (12.0%) | 525 (10.4%) | 305 (11.4%) | 90 (11.4%) | 133480 (15.7%) |
| 4 | 122850 (15.1%) | 2525 (9.3%) | 340 (7.9%) | 215 (8.0%) | 55 (6.6%) | 126040 (14.9%) |
| 5 | 92255 (11.4%) | 1570 (5.8%) | 195 (3.9%) | 95 (3.5%) | 35 (4.2%) | 94150 (11.1%) |
| Missing | 48093 (5.9%) | 2525 (9.3%) | 605 (12.0%) | 365 (13.7%) | 145 (17.9%) | 51735 (6.1%) |
| **Age** |  |  |  |  |  |  |
| Median | 36 | 31 | 32 | 33 | 39 | 36 |
| 25th Centile | 26 | 24 | 24 | 25 | 29 | 26 |
| 75th Centile | 50 | 43 | 46 | 48 | 52 | 50 |
| **Gender** |  |  |  |  |  |  |
| Female | 463755 (57.1%) | 18045 (66.1%) | 3350 (66.2%) | 1675 (62.5%) | 475 (59.4%) | 487300 (57.5%) |
| Male | 347925 (42.9%) | 9235 (33.8%) | 1705 (33.8%) | 1005 (37.5%) | 325 (40.4%) | 360195 (42.5%) |
| Unspecified | 35 (0.0%) | - | - | - | - | 35 (0.0%) |
| Missing | 25 (0.0%) |  |  |  |  | 25 (0.0%) |
|  | (N=811740) | (N=27285) | (N=5055) | (N=2675) | (N=800) | (N=847555) |
| **Total Contacts** | 1152015 | 160835 | 51170 | 43620 | 35005 | 1442645 |
| **Contact days per person** | 1-4 | 5-8 | 9-12 | 13-24 | 25+ | Overall |
|  | (N=372795) | (N=11170) | (N=2665) | (N=2020) | (N=895) | (N=389550) |
| **IMD Quintile** |  |  |  |  |  |  |
| 1 | 116895 (31.4%) | 5160 (46.2%) | 1240 (46.5%) | 915 (45.3%) | 390 (43.4%) | 124600 (32.0%) |
| 2 | 51255 (13.7%) | 1815 (16.3%) | 425 (15.9%) | 325 (16.0%) | 150 (16.5%) | 53965 (13.9%) |
| 3 | 40275 (10.8%) | 1170 (10.5%) | 290 (11.0%) | 205 (10.2%) | 90 (10.1%) | 42040 (10.8%) |
| 4 | 36495 (9.8%) | 940 (8.4%) | 195 (7.3%) | 145 (7.3%) | 65 (7.5%) | 37845 (9.7%) |
| 5 | 25470 (6.8%) | 460 (4.1%) | 90 (3.3%) | 75 (3.7%) | 25 (3.0%) | 26120 (6.7%) |
| Missing | 102405 (27.5%) | 1625 (14.5%) | 425 (15.9%) | 355 (17.6%) | 175 (19.5%) | 104980 (26.9%) |
| **Age** |  |  |  |  |  |  |
| Median | 41 | 45 | 45 | 42 | 40 | 42 |
| 25th Centile | 30 | 31 | 32 | 30 | 29 | 30 |
| 75th Centile | 54 | 56 | 55 | 53 | 51 | 54 |
| **Gender** |  |  |  |  |  |  |
| Femaile | 187600 (50.3%) | 5815 (52.1%) | 1360 (51.0%) | 1020 (50.4%) | 460 (51.5%) | 196255 (50.4%) |
| Male | 179675 (48.2%) | 5310 (47.5%) | 1295 (48.5%) | 995 (49.3%) | 430 (47.7%) | 187700 (48.2%) |
| Unspecified | 2110 (0.6%) | 45 (0.4%) | 15 (0.5%) |  |  | 2185 (0.6%) |
| Missing | 3410 (0.9%) |  |  |  |  | 3410 (0.9%) |
|  | (N=372795) | (N=11170) | (N=2665) | (N=2020) | (N=895) | (N=389550) |
| **Total Contacts** | 486260 | 66875 | 27095 | 34030 | 41085 | 655345 |

### Distribution of contacts per person

As hypothesised, the distribution of contacts per person was heavy-tailed and approximated to a power law. Figure 1 shows the logarithmic plots of the conditional cumulative distribution function for both NHS 111 and ambulance dispatch contacts with the data points forming a straight line between 5 and approximately 100 contacts.

**Figure 1.**
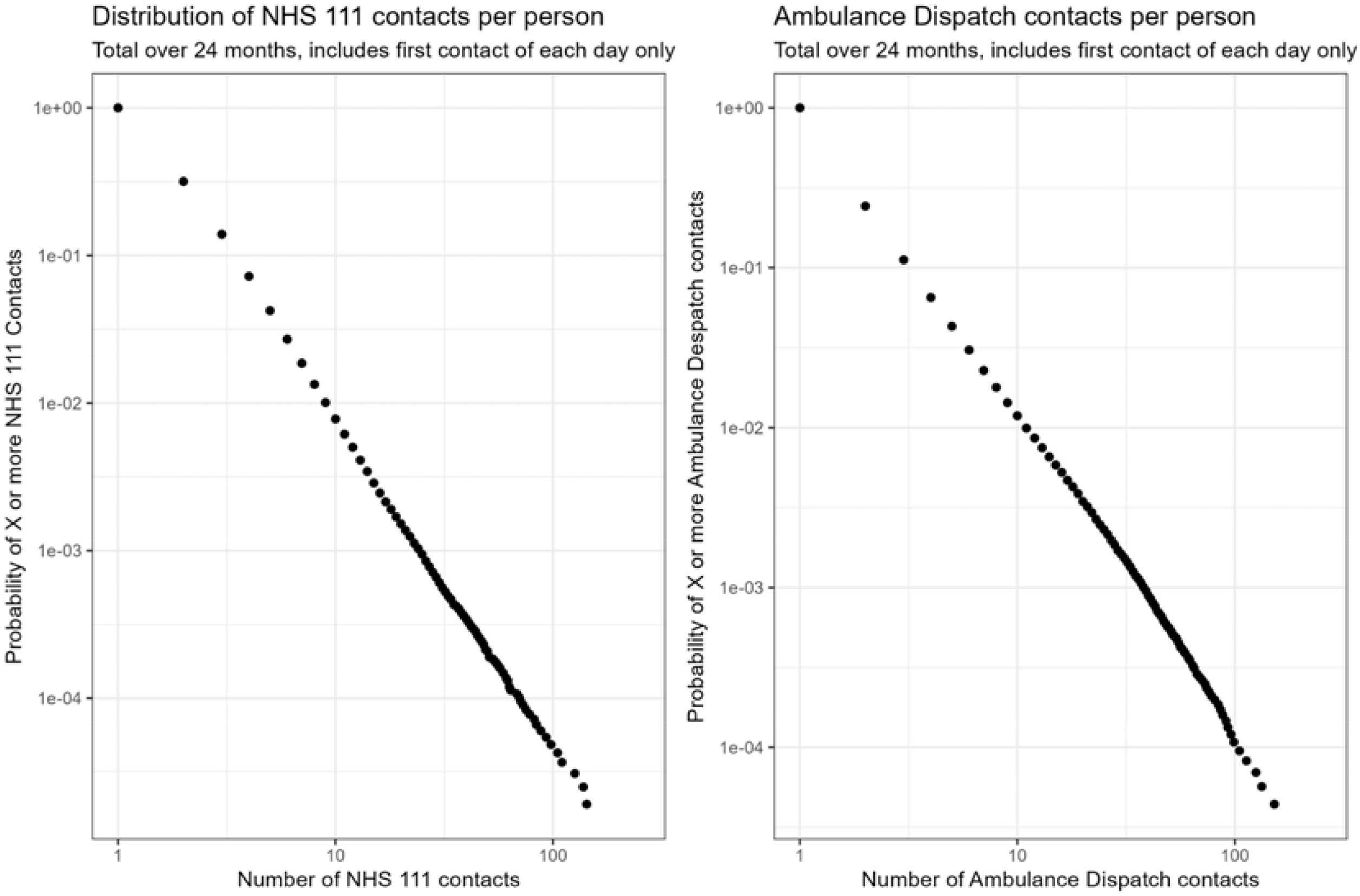
Distribution of contacts per person

Statistical fitting of the heavy tailed distributions showed that for a minimum inclusion value of 5 contacts, in the NHS 111 data a power law distribution fitted much better than a poisson distribution (p <10^−16^) and marginally better than a log-normal (p = 0.041). For the ambulance dispatch data, a power law distribution fitted much better than a poisson distribution (p <10^−16^) and marginally less well than a log-normal distribution (p = 0.024). Power law coefficients were 3.27 (5% confidence interval 3.25 to 3.29) for NHS 111 data and 2.74 (2.72 to 2.77) for the ambulance dispatch data.

### Characteristics of higher users

Table 1 summarises the characteristics of individuals in each of the five attendance groups. Supplementary figure 1 shows the same data as in Figure 1 but split by median age by sex and by IMD quintile. Table 2 summarises the results of the logistic regression. Together these indicate that while age and living in an area with the most deprived quintile of IMD were associated with having 5 or more contacts in those with at least one, they had very small or negligible effects on predicting higher levels of use. While having a contact for cardiopulmonary or neurological disorders was associated with having 5 or more contacts compared to less, beyond this (e.g. 8 or more contacts in people with at least 5) the effect was reversed and the presence of either of these made ongoing high use less likely. On the other hand having a mental health reason for contact in one or more prior contacts was predictive of ongoing use across all levels of use.

**Table 2.**
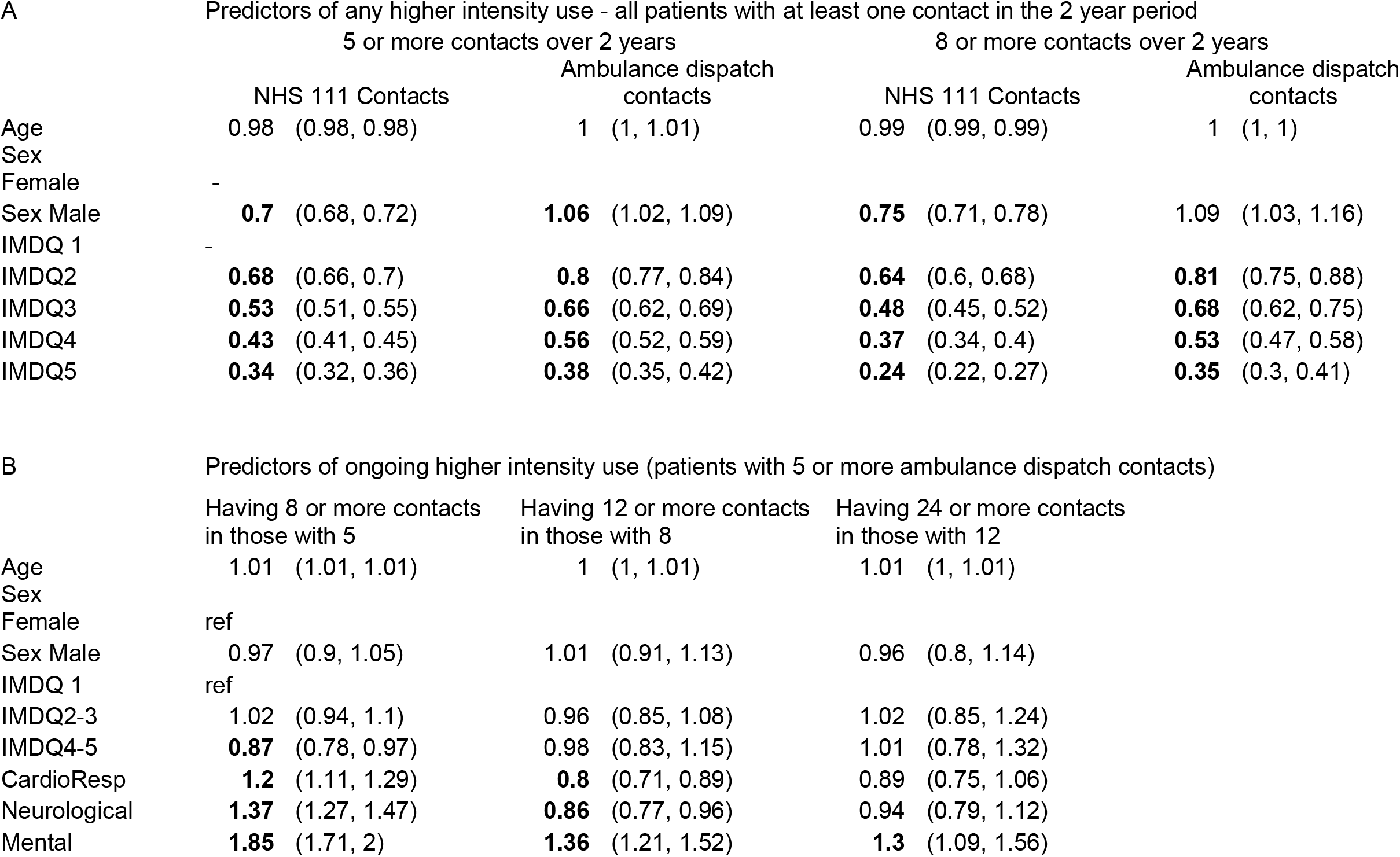
Odds ratios for predictors of higher intensity use.

### Stability of high user status

Figure 2 shows alluvial plots for distribution of attendance per person over the three 8-month time periods. These indicate that approximately 60% of high users in one period are no longer high users in the next, independently of the threshold used for high use. Supplementary figure 2 depicts the distribution of attendance at the population level over the same three periods and shows it to be remarkable stable, despite the high individual instability.

**Figure 2.**
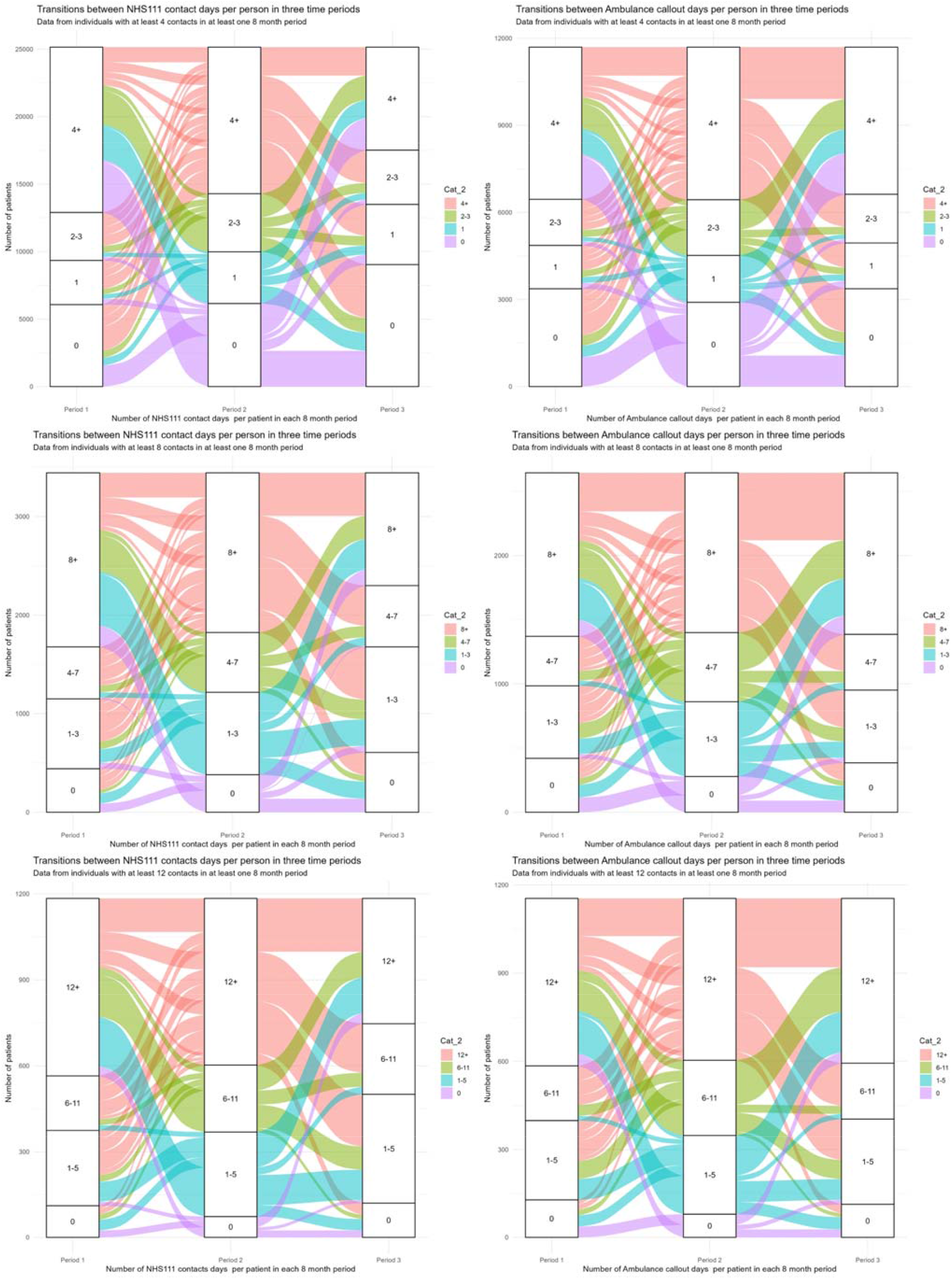
Alluvial plots showing transitions between user states

### Distribution of burst lengths

Figure 3 shows the distribution of burst length using three different time windows. For each time window, and for both services, the distributions are clearly heavy tailed. However while the NHS 111 contacts distribution has the characteristic appearance of a power law, the ambulance dispatch data is convex in the tail, decaying faster than would be the case of a power law. This may be explainable by interventions for high intensity use being triggered by sustained patterns of activity resulting in action to reduce use.

**Figure 3.**
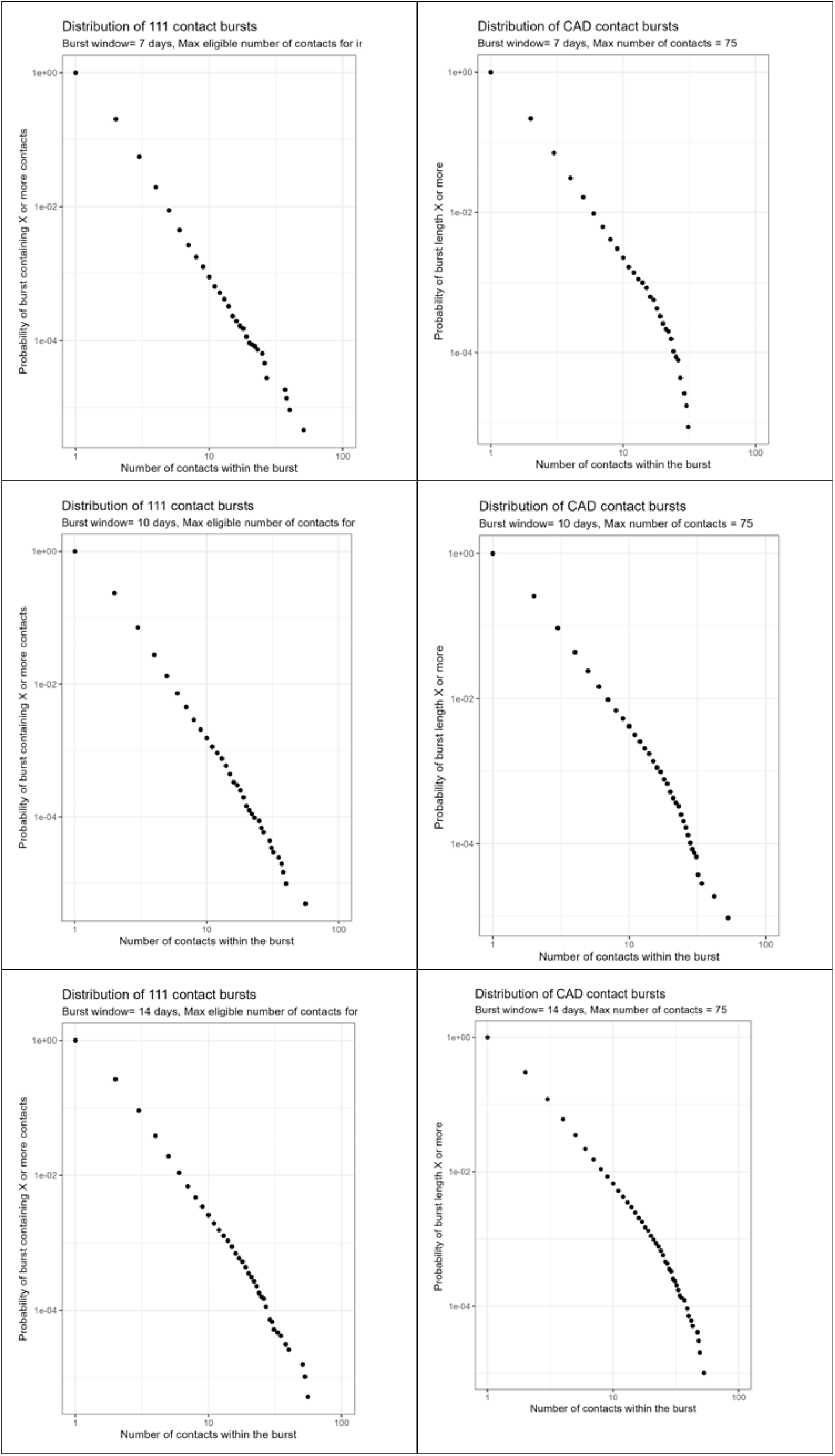
Distribution of lengths of bursts of contacts.

## Discussion

### Summary of main findings

High use of pre-hospital emergency medical services shows multiple features indicative of a complex system. The distribution of contacts per individual is continuous, heavy-tailed and approximates to a power law. Contacts by an individual are linked in time as evidenced by both bursty patterns of contact and of the instability of high user status over multiple time periods. While age and socioeconomic deprivation are associated with moderately high use compared to no use, contacts for mental health reasons are the factor most strongly associated with ongoing high intensity use.

### Strengths and limitations

This is the first study examining patterns of pre-hospital emergency care use from a complex systems perspective. The database was large (Yorkshire Ambulance Service provides prehospital emergency care for a population of approximately 5 Million people) and has previously been validated. The statistical methods had previously been used to study emergency department and primary care out of hours services and analysis was carried out independently by all three authors, with differences in code and results reviewed and discussed. The main limitation was the data regarding reasons for contact which used broadly defined categories representing the pragmatic nature of this data collection. These included the source of referral rather than the patient’s reason for contact in 17.2% of contacts and the non-specific category “other” in 15.4% of contacts. Furthermore we limited contacts to the first of each day – while it is possible that some individuals may have had multiple discrete events we were unable to distinguish between these and follow-up calls for instance reporting a change in the clinical situation. We set our inclusion threshold to a deliberately low level of 5 or more contacts within a 2 year period. This is a smaller number over a longer time period than current thresholds for case-management interventions (such as 5 contacts in a month or 12 over a three month period) but enables very high users to be seen in the context of a system-wide phenomenon which is applicable to the top 5% of prehospital emergency service users. Importantly there was no change in the shape of the distribution at higher intensity of use (e.g. between 50 and 100+ contacts over the 2 year period), suggesting common underlying mechanisms in those with different patterns of use.

### Implications for practice, policy and research

The finding of multiple features of complex systems in pre-hospital emergency care show strong similarities with those for high intensity emergency department attendance and primary care out of hours service use. These findings are compatible with a complex generative model for high intensity in which social interaction mechanisms drive high use. These findings present insights for addressing the challenge of high intensity use. This evidence of system-level effects points towards the importance of a multifactorial approach that goes beyond simply identifying and targeting individual high intensity users. Importantly, the high level of temporal instability at the individual level means that many interventions that appear to work may actually be observing natural variation. The consistency of the findings across all levels of use, in particular the presence of bursts, suggests that simply waiting for high intensity users to reach a given threshold of contacts before intervention may be less effective than identifying patterns of high-risk contact behaviour and intervening on this. The association of mental health issues with ongoing high intensity use means that measures addressing this are likely to be at least one part of any intervention.

## Conclusion

Prehospital emergency service use shows multiple statistical features typical of a complex system. Interventions to manage demand need to consider both individual high intensity users (particularly in relation to their mental health) and the behaviour of the whole system.

## Data Availability

Original data analysed for this study is securely held by the DataConnect Secure Data Environment at the University of Sheffield. Analysis scripts are available upon reasonable request to the authors

## Acknowledgement

We gratefully acknowledge the University of Sheffield Data Connect Service, which provided data management support and the data extract from the CUREd+ Research Database, and the University of Sheffield Secure Data Services who hosted the data in the secure data environment. This work uses data provided by patients and collected by the NHS as part of their care and support. Funding for the data preparation of the CUREd+ Research Database was funded in part by the Connected Cities study, now continued through the NIHR Applied Research Collaboration and the NHS England Yorkshire & Humber Secure Data Environment. The views expressed in this paper are those of the authors and do not represent those of The University of Sheffield, NHSA, NHS, NIHR or the Department of Health and Social Care.

## CREDIT Taxonomy

Cussens: Formal Analysis, Writing (Drafting);

Do: Formal Analysis, Writing (Drafting);

Chambers: Data Curation; Writing (review and editing)

Crum: Methodology; Writing (review and editing)

Burton Conceptualisation, Methodology, Supervision, Formal Analysis, Writing (review and editing)

