## Supplementary Data for "Statistical features of complex systems in use of pre-hospital emergency services: a linked database study"

Patterns of high intensity use of pre-hospital emergency services show multiple features of complex systems.

Supplementary Data

Supplementary Table 1 Reason for contact with Ambulance Dispatch Service in high users grouped by high intensity use category

| Number of contacts per individual | 5-8 | | 9-12 | | 13-24 | | 25+ | |
| --- | --- | --- | --- | --- | --- | --- | --- | --- |
| CardioRespiratory | 12040 | 18.0% | 4850 | 17.9% | 5980 | 17.6% | 7600 | 18.5% |
| Musculoskeletal | 2725 | 4.1% | 1260 | 4.7% | 1370 | 4.0% | 1540 | 3.7% |
| Mental | 7720 | 11.5% | 3480 | 12.8% | 5600 | 16.5% | 9270 | 22.6% |
| Neurological | 8575 | 12.8% | 3665 | 13.5% | 4540 | 13.3% | 4825 | 11.7% |
| Trauma | 3660 | 5.5% | 1390 | 5.1% | 1750 | 5.1% | 1750 | 4.3% |
| Infection | 3695 | 5.5% | 1550 | 5.7% | 1820 | 5.3% | 2480 | 6.0% |
| GI/GU | 1510 | 2.3% | 580 | 2.1% | 695 | 2.0% | 855 | 2.1% |
| Sexual/Reproductive | 380 | 0.6% | 100 | 0.4% | 70 | 0.2% | 75 | 0.2% |
| General | 2855 | 4.3% | 1310 | 4.8% | 1455 | 4.3% | 1660 | 4.0% |
| Other | 10280 | 15.4% | 4120 | 15.2% | 5090 | 15.0% | 5600 | 13.6% |
| Internal Referral | 8775 | 13.1% | 3065 | 11.3% | 3900 | 11.5% | 4250 | 10.3% |
| Health Professional | 4575 | 6.8% | 1705 | 6.3% | 1730 | 5.1% | 1140 | 2.8% |
| Uncoded | 55 | 0.1% | 20 | 0.1% | 30 | 0.1% | 25 | 0.1% |

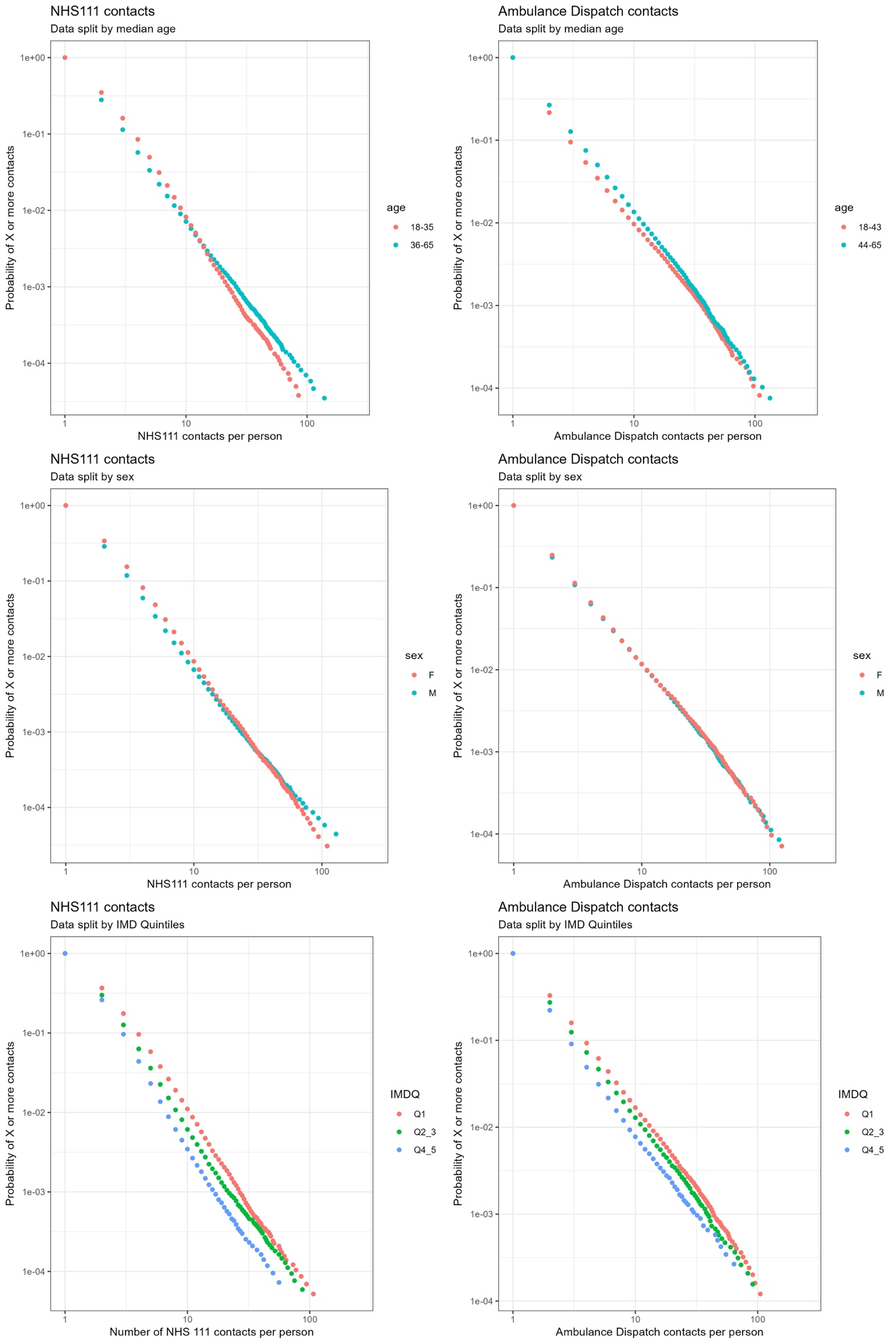

Supplementary Figure 1 Distribution of contacts per person, split by median age, sex and IMD category

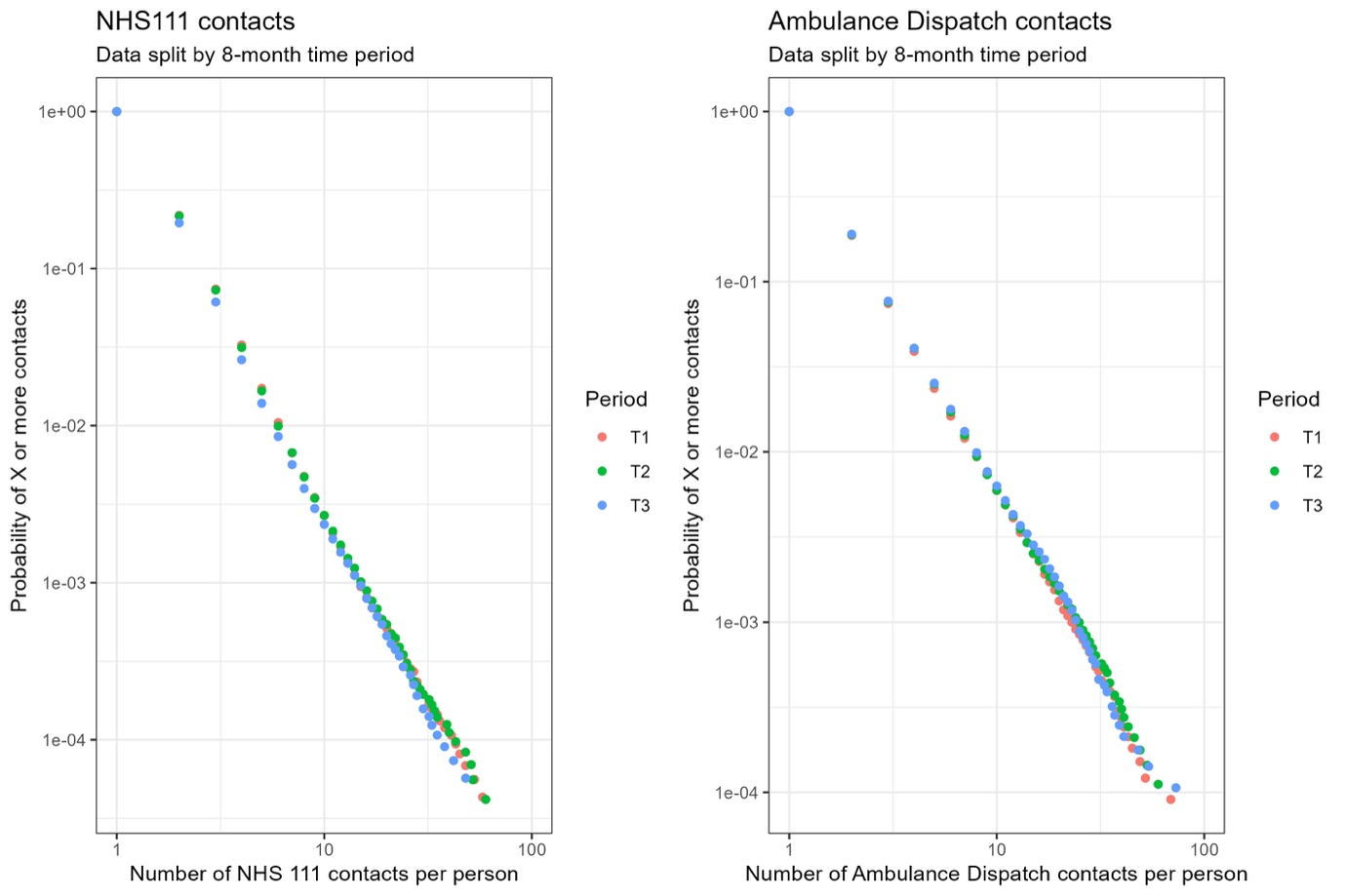

Supplementary Figure 2 Distribution of contacts per person over 3 consecutive 8-month time periods.
